# Novel plasma biomarkers of amyloid plaque pathology and cortical thickness: evaluation of the NULISA targeted proteomic platform in an ethnically diverse cohort

**DOI:** 10.1101/2024.12.07.24318660

**Authors:** Xuemei Zeng, Anuradha Sehrawat, Tara K. Lafferty, Yijun Chen, Mahika Rawat, M. Ilyas Kamboh, Victor L. Villemagne, Oscar L. Lopez, Ann D. Cohen, Thomas K. Karikari

## Abstract

**INTRODUCTION:** Proteomic evaluation of plasma samples could accelerate the identification of novel Alzheimer’s disease (AD) biomarkers. We evaluated the novel NUcleic acid Linked Immuno-Sandwich Assay (NULISA^TM^) proteomic method in an ethnically diverse cohort.

**METHODS:** Plasma biomarkers were measured with NULISA in the Human Connectome Project, a predominantly preclinical biracial community cohort in southwestern Pennsylvania. Selected biomarkers were additionally measured using Simoa and Quest immunoassays.

**RESULTS:** On NULISA, phosphorylated tau (p-tau217, p-tau231, p-tau181), GFAP, and MAPT-tau showed the top significant association with Aβ PET status, followed by neuroinflammation markers CCL2, CHIT1, CXCL8, and the synaptic marker NRGN. Biomarkers associated with cortical thickness included astrocytic protein CHI3L1, cytokine CD40LG, growth factor BDNF, Aβ-associated metalloprotein TIMP3, and FCN2 linked with brain atrophy in AD. Furthermore, moderate to strong between-platform correlations were observed for various assays.

**DISCUSSION:** NULISA multiplexing advantage allowed concurrent assessment of established and novel plasma biomarkers of Aβ pathology and neurodegeneration.

## 1. BACKGROUND

Robust blood-based biomarkers will greatly enhance the clinical management of Alzheimer’s disease (AD), aiding risk assessment, diagnosis, staging, and therapy monitoring.^1–3^ Traditionally, a definitive AD diagnosis requires post-mortem confirmation of amyloid-beta (Aβ) plaques and tau neurofibrillary tangles in the brain.^4,5^ However, clinically approved cerebrospinal fluid (CSF) biomarkers and brain imaging techniques, such as magnetic resonance imaging (MRI) and positron emission tomography (PET) scans, now support the antemortem diagnosis of AD.^6–9^ Despite their effectiveness, these modalities are expensive and relatively invasive, making them unsuitable for large-scale population use.

In recent years, numerous technological advancements have enabled significant transformation in AD biomarker research. These innovations have enhanced the sensitivity, precision, and overall performance of blood-based AD biomarkers. Various automated and semi-automated platforms, such as Single molecule array (Simoa), Meso Scale Discovery (MSD), Immunomagnetic Reduction (MagQu), and Lumipulse electrochemiluminescence (ECL) immunoassays, are now available for measuring AD biomarkers in blood.^10^ These biomarkers, including Aβ peptides, multiple phosphorylated tau (p-tau) species, glial fibrillary acidic protein GFAP, neurofilament light chain (NEFL) and brain-derived tau (BD-tau), offer cost-effective ways to gain critical insights into AD-associated brain pathologies.^2,11–15^ However, the current blood biomarkers have primarily focused on classical AD pathologies such as Aβ plaques (A), neurofibrillary tangles (T), and markers of neurodegeneration (N).

AD is a complex, multifactorial disease with various pathophysiological processes. In addition to AT(N), several other processes, such as inflammation (I), vascular dysregulation (V), synucleinopathy (S), and synaptic malfunction, have been linked to AD pathogenesis.^16–20^ This complexity underscores the need for a more comprehensive panel of purposely selected biomarkers to capture the complex interplay among these pathologies. Notably, a multiplex Nucleic acid-Linked Immuno-Sandwich Assay (NULISA) panel, specifically the NULISAseq CNS disease panel, was recently introduced^21^. This panel utilizes NULISA, a novel automatic proteomic platform, to simultaneously profile ∼120 proteins associated with a broad spectrum of neurodegenerative disorders. NULISA, built as an extension of proximity ligation assay (PLA) technology, integrates multiple mechanisms to enhance the performance of PLA, including a proprietary sequential immunocomplex capture and release mechanism for background reduction, next-generation sequencing (NGS)-based signal readout, and fine-tuning the ratio of unconjugated “cold” antibodies to DNA-conjugated “hot” antibodies to mitigate sequencing reads of high-abundant proteins. It can detect hundreds of proteins with attomolar sensitivity and ultrabroad dynamic range.^21^

Utilizing only 10 μl of plasma, the NULISAseq CNS panel profiles several classical AD biomarkers, including MAPT-tau, multiple phosphorylated tau species (p-tau181, p-tau217, and p-tau231), Aβ peptides (Aβ38, Aβ40, and Aβ42), GFAP, and NEFL. Additionally, it measures key proteins involved in various brain pathophysiological processes, such as α-synuclein for synucleinopathy, ICAM1, VCAM1, and VEGFs for vascular function, microglial biomarkers TREM1 and TREM2, astrocytic biomarkers CHI3L1 and GFAP, and synaptic markers like NPTX and NRGN.

We previously applied the NULISAseq CNS panel to a population-based cohort of predominantly cognitively normal older non-Hispanic White individuals.^22^ The results showed good alignment between NULISAseq outcomes and those obtained with Simoa-based assays for several classical AD biomarkers. Additionally, we identified novel plasma biomarkers that showed significant associations with neuroimaging-quantified A, T and N ground truth biomarkers cross-sectionally and/or longitudinally. Consistent with our findings, Ibanez et al. also reported good alignment between NULISAseq measurements and several other validated assays, such as Simoa, Lumipulse, Single Molecule Counting technology, and immunoprecipitation-mass spectrometry assays.^23^ While these earlier investigations support using the NULISAseq CNS panel to provide comprehensive characterization of AD pathologies, further evaluation is still needed.

The primary focus of this study was to assess the performance of the NULISAseq CNS Disease Panel using a different community-based cohort, which is racially more diverse and consists of a mix of Black/African Americans and non-Hispanic Whites. We evaluated the association between NULISAseq biomarkers with Aβ pathology and neurodegeneration statuses defined by Aβ PET and cortical thickness, respectively. Additionally, we assessed associations between NULISAseq biomarkers and common AD risk factors such as age, *APOE* genotype, self-identified race, and sex.

## 2. METHODS

### 2.1 Participants

The study cohort consisted of participants from the Human Connectome Project (HCP) in Pittsburgh, Pennsylvania, USA. HCP is a community-based study aimed at improving the understanding of the relationships between AD and cerebrovascular disease.^24^ Participants aged 50–89 were recruited primarily through the University of Pittsburgh Alzheimer’s Disease Research Center and the Pitt + Me web portal. All participants provided written consent, and the University of Pittsburgh Institutional Review Board approved the study.

Comprehensive demographic, behavioral, and laboratory data were collected from participants upon enrollment. Each participant underwent neurophysiological assessment, brain structural and functional imaging (functional magnetic resonance imaging [MRI], magnetoencephalography [MEG]), and [^11^C] Pittsburgh Compound B (PiB) PET imaging over a three-day period. Blood was collected using EDTA-containing tubes before the imaging and centrifuged at 2000xg for 10 minutes at 4°C to separate plasma from blood cells, following standard guidelines.^10^ Plasma was aliquoted into low-bind polypropylene tubes and stored at −80°C until analysis. Buffy coat was collected for *APOE* genotyping as previously described.^25^ Participants with one or two ε4 alleles were classified as *APOE* ε4 carriers and others as non-carriers. Participants’ Aβ pathology (A) and neurodegeneration (N) statuses were classified according to [^11^C] PiB PET (global standardized uptake value ratio [SUVR] >1.346 as A+) and MRI scans for cortical thickness (a surface-area weighted average cortical thickness of regions of interest < 2.7 as N+), as previously described.^26,27^ Detailed study design for the HCP study, including recruitment strategies, multi-domain cognitive assessments, neuroimaging, and data processing, can be found in a previous publication.^24^

### 2.2 NULISAseq CNS Disease Panel 120 assay

Alamar Biosciences, Inc. conducted the NULISAseq CNS Disease Panel 120 assay on an Alamar ARGO^TM^ prototype system following published protocols. ^21,22^ Briefly, thawed plasma samples were centrifuged at 10,000xg for 10 minutes to remove particulates, then incubated with a cocktail of capture and detection antibodies linked to DNA barcodes. The resulting immunocomplexes were purified, and a ligation mix containing T4 DNA ligase and a specific DNA ligator sequence was used to generate cDNA sequences by ligating the DNA barcodes of capture and detection antibody pairs. These reporter DNA levels were then measured using Next-Generation Sequencing (NGS). All samples were analyzed in the same run, with two replicates of a sample control (SC), three replicates of an inter-plate control (IPC), and two replicates of a negative control included to monitor assay performance. NULISA Protein Quantification (NPQ) was derived by dividing target counts by internal control counts of each well and then by the median IPC counts, followed by log2-transformation. Fold changes between groups were calculated as 2 to the power of the difference in NPQ. The average CV of the duplicate measure of SC across 116 biomarkers was 5.5%.

### 2.3 Procedures for other immunoassay platforms

Simoa assays were performed on an HD-X instrument (Quanterix, Billerica, MA, USA) using commercialized kits. Thawed plasma samples were centrifuged at 4000xg for 10 minutes to remove particulates before the analysis. NEFL, GFAP, Aβ42, and Aβ40 were measured using the Neurology 4-Plex E kit (#103670). The p-tau181 levels were assessed with the p-tau181 V2 Advantage kit (#103714), and the p-tau217 levels were measured using the ALZpath Simoa® p-Tau 217 V2 Assay Kit (#104371). IL6 and TNF were also measured using immunoassays available from Quest Diagnostics for clinical blood tests.^28,29^

### 2.4 Statistical analysis

Data analysis was performed using MATLAB (version R2021b). For demographic characteristics, the Wilcoxon rank-sum test was used for two-group comparisons of continuous variables, while Fisher’s exact test was applied to categorical variables. The Wilcoxon rank-sum test was also used to assess the significance of the association between NULISAseq biomarker levels and A, N, *APOE* ε4 carrier, and self-identified racial statuses. Spearman’s rank correlation was employed to evaluate the strength and direction of associations between two continuous variables. False discovery rates (FDR) were calculated using the procedure Yoav Benjamini and Yosef Hochberg described in 1995.^30^ A *p*-value < 0.05 and FDR < 5% were considered significant for all comparisons, while a *p*-value < 0.05 but FDR > 5% was considered marginally significant. A *p*-value > 0.05 was considered non-significant. Receiver operating characteristic (ROC) curves and the area under the curve (AUC) were based on the generalized linear regression models. Confidence intervals were estimated using bootstrap (1000 replicates).

## 3. RESULTS

### 3.1 Participant characteristics

The study cohort included 88 participants from the HCP study ^24^. The median age was 70 years (IQR 9.5), with 41 (46.6 %) females, 32 (36.4%) participants whose self-identified race/ethnicity was not non-Hispanic White (31 African American and 1 Asian), and 31 *APOE* ε4 carriers (35.2%) (see Table 1 for demographic characteristics). All except one participant had completed at least 12 years of education. Most participants were cognitively normal at the time of study enrollment, with 65 (73.9%) having Montreal Cognitive Assessment (MoCA) scores >= 24. Participants were classified based on ^11^C-PiB Aβ PET status (60 A- and 28 A+) and MRI-based neurodegeneration status (53 N- and 35 N+). Aβ PET positivity was associated with older age (*p* = 0.001) and higher education (15 [IQR=6] years versus 16 [IQR=4] years, *p* = 0.025). Additionally, a higher A+ rate was observed in non-Hispanic White participants compared to others (all except one being African American), with 30.3% and 19.4% being A+, respectively. *APOE* ε4 carriers had a slightly higher likelihood of being N+ (54.8% in carriers compared to 31.6% in non-carriers, *p* = 0.042).

**Table 1:** Characteristics of cohort participants. The median and interquartile range (IQR) are reported for continuous variables, while frequencies are shown for categorical variables. *All except one non-Hispanic White were African American. ^#^P-values were calculated using the Wilcoxon rank-sum test for continuous variables and Fisher’s exact test for categorical variables. Aβ PET status is based on the global PiB SUVR, with > 1.346 indicating Aβ+. N status is based on cortical thickness assessed by structural MRI, with thickness < 2.7 indicating positive status.

| | | By Aβ PET status | | | By neurodegeneration status | | |
| --- | --- | --- | --- | --- | --- | --- | --- |
| | Total | Aβ- | Aβ+ | *p*-value<sup>#</sup> | N- | N+ | *p*-value<sup>#</sup> |
| **N (%)** | 88 | 60 (68.1%) | 28 (31.8%) |  | 53 (60.2%) | 35 (39.8%) |  |
| **Age (median, IQR)** | 70 (9.5) | 67.5 (12) | 73.3 (10) | 0.001 | 70 (8) | 69 (11.8) | 0.717 |
| **Sex (n, %)** |  |  |  | 0.370 |  |  | 0.385 |
| **Female** | 41 | 30 (73.2%) | 11 (26.8%) |  | 27 (65.9%) | 14 (34.1%) |  |
| **Male** | 47 | 30 (63.8) | 17 (36.2%) |  | 26 (55.3%) | 21 (44.7%) |  |
| **Years of education (mean, SD)** | 16 (5) | 15 (6) | 16 (4) | 0.025 | 16 (4) | 14 (6) | 0.171 |
| **Self-identified race (n, %)** |  |  |  | 0.027 |  |  | 0.106 |
| **Non-Hispanic White** | 56 | 34 (60.7%) | 22 (39.3%) |  | 36 (64.3%) | 20 (35.7%) |  |
| **Other<sup>\*</sup>** | 32 | 26 (80.6%) | 6 (19.4%) |  | 17 (54.8%) | 15 (45.2%) |  |
| **MoCA (n, %)** |  |  |  | 0.190 |  |  | 0.073 |
| **$\geq 24$** | 65 | 45 (69.2%) | 20 (30.8%) |  | 42 (64.6%) | 23 (35.4%) |  |
| **<24** | 23 | 15 (65.2%) | 8 (34.8%) |  | 11 (47.8%) | 12 (52.2%) |  |
| **APOE $\epsilon 4$ carrier (n, %)** |  |  |  | 0.636 |  |  | 0.042 |
| **No** | 57 | 40 (70.2%) | 17 (29.8%) |  | 39 (68.4%) | 18 (31.6%) |  |
| **Yes** | 31 | 20 (64.5%) | 11 (35.5%) |  | 14 (45.2%) | 17 (54.8%) |  |

### 3.2 Comparison between NULISAseq and other assay platforms

We evaluated the agreement between NULISAseq measurements and those from other assay platforms, including Simoa and Quest immunoassays, for several biomarkers. These included p-tau217, p-tau181, p-tau231, GFAP, NEFL, Aβ40, and Aβ42, which were measured by both NULISAseq and Simoa assays, and TNF and IL6, measured by NULISAseq, Simoa, and Quest assays. NULISAseq measured all these biomarkers with high repeatability. The intra-plate CVs based on duplicate measurements of a sample control were 3.2% for p-tau217, 0.6% for p-tau181, 3.3% for p-tau231, 5.2% for GFAP, 0.5% for NEFL, 1.2% for Aβ40, 3.6% for Aβ42, 2.7% for TNF, and 2.5% for IL6. Spearman’s rank correlation coefficient (*rho*) was used to quantify the strength of the correlation between the platforms. The *rho* values for the correlation between NULISAseq and Simoa assays ranged from 0.357 to 0.792, as shown in Figure 1A. Among the biomarkers, GFAP and NEFL exhibited the highest correlations between NULISAseq and Simoa results, with *rho* values of 0.792 and 0.752, respectively. The correlation, although significant, was lower for Aβ peptides, with *rho* values of 0.397 and 0.466 for Aβ40 and Aβ42, respectively. The lowest correlation was observed between NULISAseq TNF and Simoa TNF, with a *rho* value of 0.357, although it is unclear if this can be explained by the smaller sample size.

**Figure 1.**
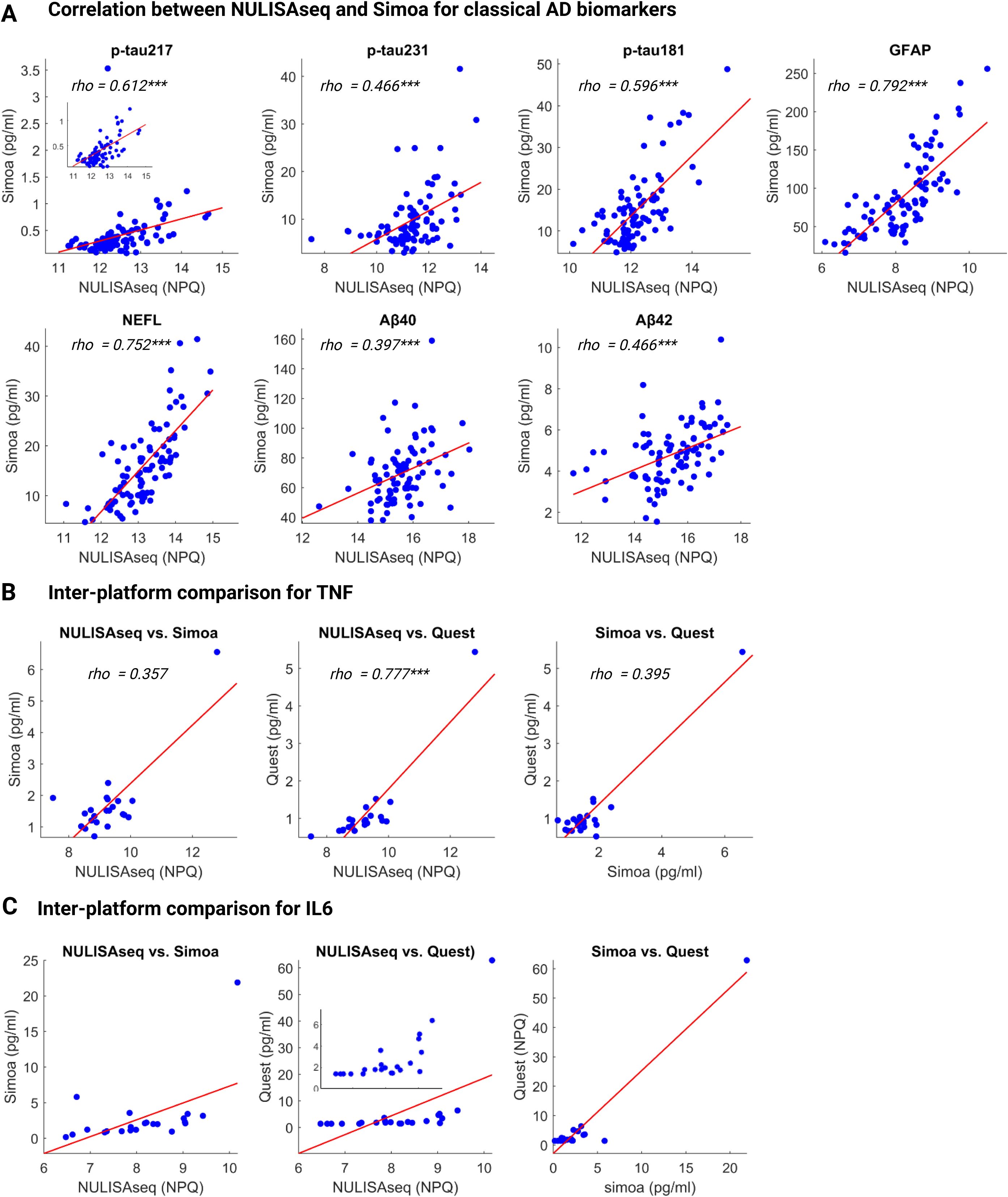
Scatter plots illustrate the inter-platform comparisons between NULISAseq and other technologies. (A) Concordance between NULISAseq and Simoa for classical AD biomarkers. (B-C) Inter-platform comparison between NULISAseq, Simoa, and Quest for TNF (B) and IL6 (C). NULISAseq biomarkers were quantified using NPQ, representing the log2-transformation of normalized target counts. The abundance units are pg/ml for Simoa and Quest immunoassays. Rho and p values were determined using Spearman rank-based correlation. Red lines indicate least squares regression lines. Statistical significance is denoted as follows: *p < 0.05, **p < 0.01, ***p < 0.001.

NULISAseq measurements showed a relatively robust correlation with results from Quest immunoassays, with *ρ* values of 0.777 and 0.748 for TNF (Figure 1B) and IL6 (Figure 1C), respectively. Notably, for both TNF and IL6, stronger correlations were observed between NULISAseq and Quest compared with between NULISAseq and Simoa (*rho* values of 0.357 for TNF and 0.586 for IL6) and between Quest and Simoa (*rho* values of 0.395 for TNF and 0.509 for IL6). In the case of TNF, neither the correlation between NULISAseq and Simoa nor Quest and Simoa was statistically significant.

### 3.3 Association between NULISAseq and A**β** pathology (A)

We utilized the Wilcoxon rank-sum test to assess the significance of the association between NULISAseq biomarkers and dichotomized Aβ pathology status, as measured by ^11^C-PiB PET imaging. Three biomarkers – p-tau217, GFAP, and p-tau231 – showed significant associations according to a 5% FDR cutoff. Consistent with the well-documented utility of p-tau217 in predicting Aβ pathology, it emerged as the most significant biomarker, exhibiting a *p*-value < 0.0001 and an average increase of 108% in A+ individuals compared with A- controls (Figure 2A). The ROC analysis utilizing the logistic regression model indicated a predictive accuracy of 0.922 (95% CI: 0.842-0.966). GFAP and p-tau231 were also increased in A+ participants, with *p*-values of 0.0004 and 0.0005 and average fold change increases of 63% and 76%, respectively. Additionally, seven biomarkers—MAPT, CCL2, p-tau181, CHIT1, CXCL8, NRGN, and NEFL—showed marginal significance, with *p*-values <0.05 but FDR >5%. All biomarkers except NRGN were increased in A+ participants (Figure 2A).

**Figure 2.**
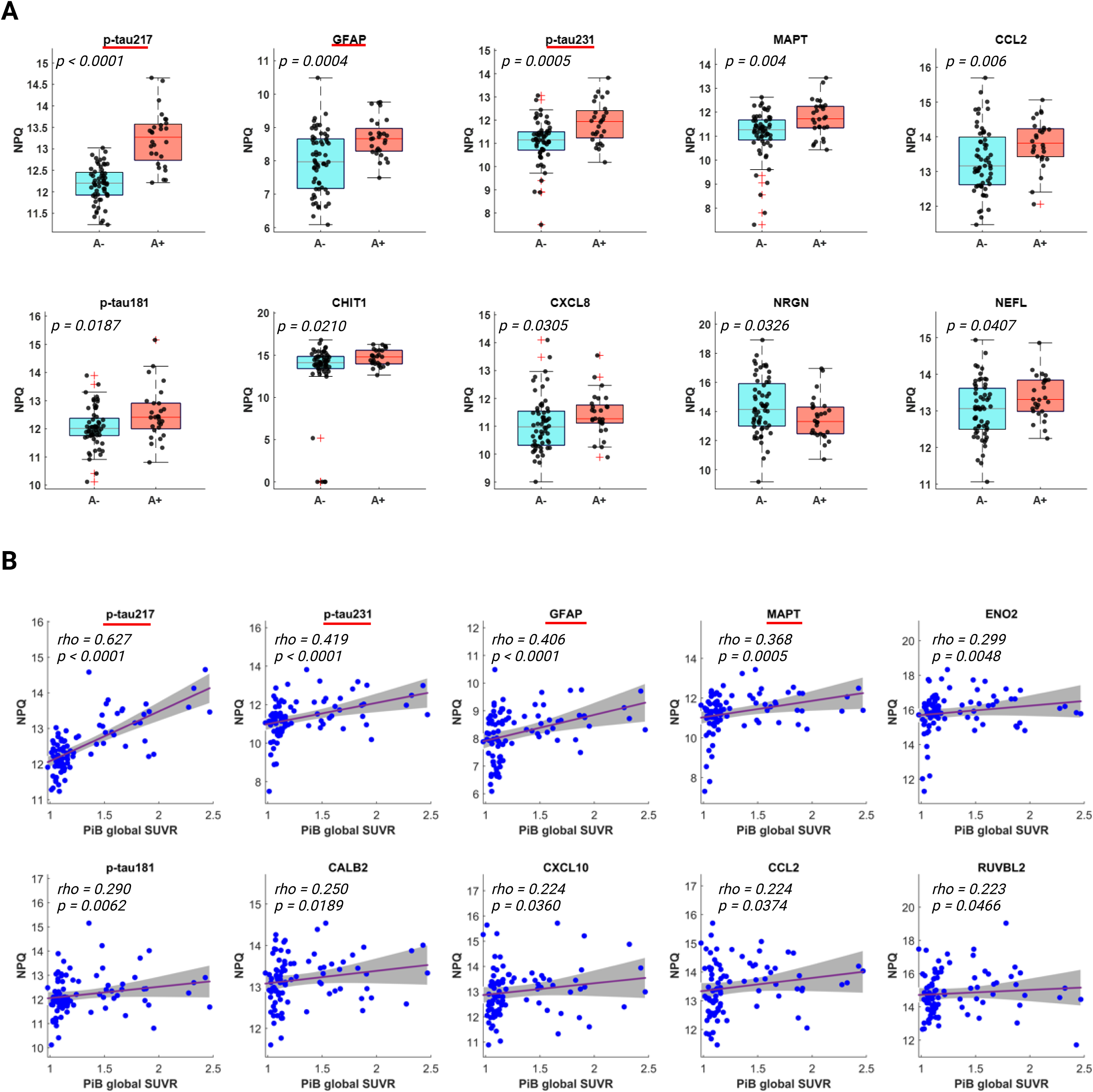
Association of NULISAseq biomarkers with Aβ PET status. (A) Box plot distributions for biomarkers with significant or marginally significant differences between Aβ PET-negative (A-) and Aβ PET-positive (A+) groups. P values were determined using Wilcoxon’s rank-sum test. Aβ PET status was determined based on [^11^C] PiB PET, with global SUVR > 1.346 as A+. (B) Scatter plots illustrating the significant correlation between biomarker levels and PiB global SUVR. The strength of the correlation was assessed using Spearman’s rank correlation. Red lines with grey bands indicate linear regression fits with 95% confidence intervals. Biomarkers with red underlines indicate having a significant association with Aβ PET positivity or PiB global SUVR according to a 5% FDR cutoff. NPQ represents the log2-transformation of normalized target counts; SUVR, Standardized Uptake Value Ratio.

To account for potential confounding effects of demographic covariates, we employed logistic regression models to assess their impact on the association between NULISAseq biomarkers and dichotomized Aβ PET status. Given the limited sample size, we included each covariate—age, sex, race, *APOE* genotype, and years of education—separately in the analysis. Among the covariates tested, age had the most significant influence on the association between NULISAseq plasma biomarkers and Aβ PET status, with GFAP and NEFL being the most affected. GFAP’s significance level changed from a *p*- value of 0.0013 (unadjusted logistic regression model) to 0.0270 when age was included as a covariate (Supplementary Table S1). NEFL, which initially showed a marginal significant association with Aβ PET status, lost its significance after adjusting for age, with a *p*-value of 0.3930. We also evaluated the significance of the association after adjusting for age, sex, and *APOE* genotype. P-tau217 remained significant, while GFAP, p-tau231, p-tau181, MAPT, and CCL2 showed marginally significant associations with Aβ PET status. CHIT1, CXCL8, NRGN, and NEFL no longer showed significant associations.

We also evaluated the association between NULISAseq biomarkers and PiB global SUVR. Four biomarkers—p-tau217, p-tau231, GFAP, and MAPT—showed significant correlations, with FDR < 5% (Figure 2B). Additionally, six biomarkers—ENO2, p-tau181, CALB2, CXCL10, CCL2, and RUVBL2— showed marginally significant correlations (*p*-values < 0.05 but FDR > 5%) (Figure 2B). All selected biomarkers showed a positive correlation with PiB SUVR.

### 3.4 Association between NULISAseq and neurodegeneration (N)

No NULISAseq biomarker was significantly associated with dichotomized N status in this cohort, as assessed by MRI-measured cortical thickness, according to the FDR cutoff of 5% using the Wilcoxon rank-sum test. However, five biomarkers—BNDF, TIMP3, FCN2, CHI3L1, and CD40LG—showed marginal significance with *p*-values <0.05 but FDR >5% (Figure 3A). Among these, BDNF had the highest significance, with a *p*-value of 0.005, and was elevated in N+ individuals, showing an average increase of 90% compared to N- controls. TIMP3, CHI3L1, and CD40LG were also increased in N+ individuals, with average increases of 73% (*p* = 0.027), 64% (*p* = 0.035), and 57% (*p* = 0.042), respectively. FCN2 had lower levels in N+ participants, with an average decrease of 15% (*p* = 0.027). BDNF remained the most significant biomarker after adjusting for age, sex, and *APOE* genotype, with a p-value of 0.014. This was followed by CHI3L1 (p = 0.015), BASP1 (p = 0.022), TIMP3 (p = 0.028), FCN2 (p = 0.041), YWHAG (p = 0.044), and CCL13 (p = 0.049).

**Figure 3.**
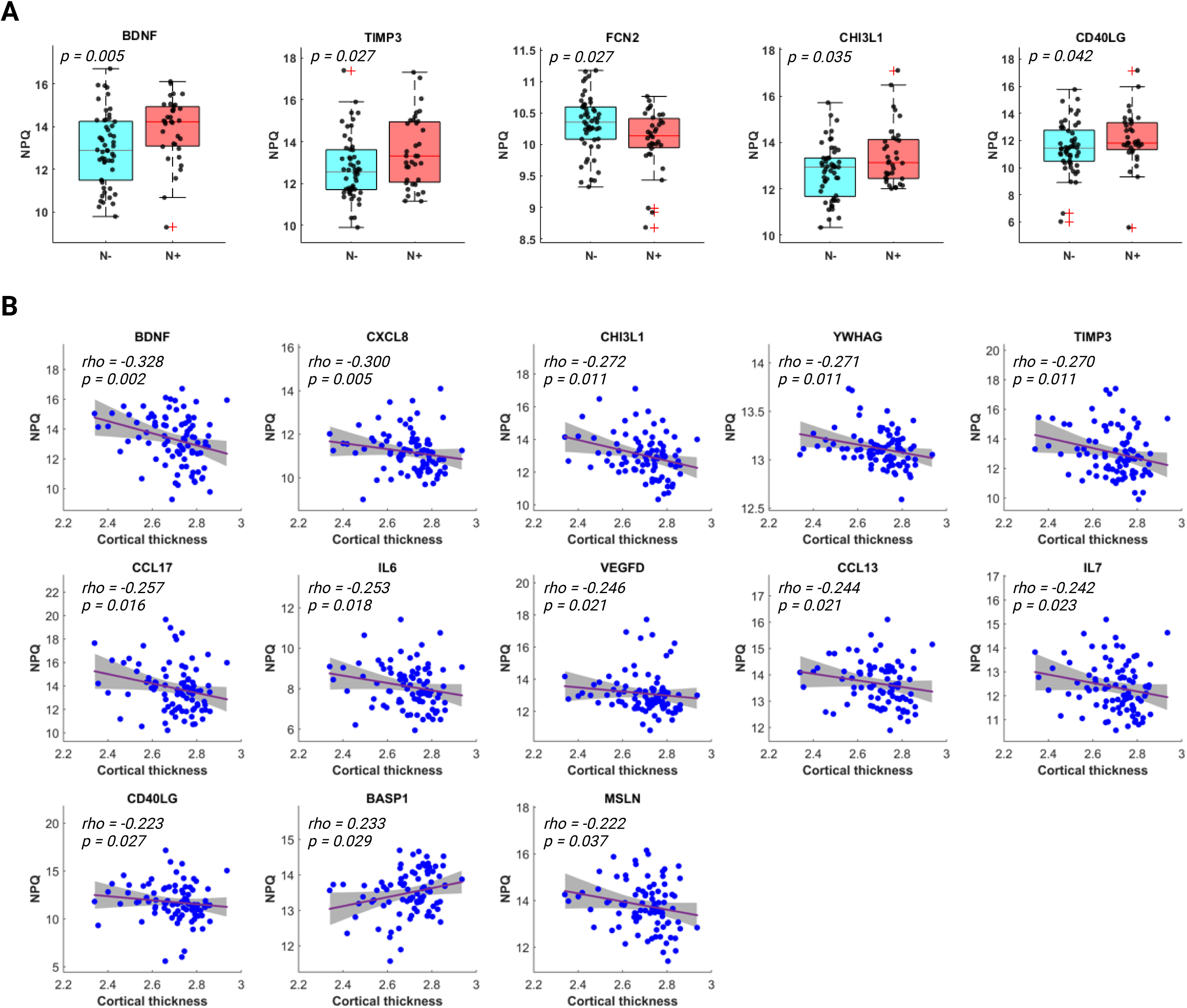
Association of NULISAseq biomarkers with neurodegeneration (N) Status. (A) Box plot distributions of biomarkers with marginal significant associations with N Status. P values were determined using Wilcoxon’s rank-sum test. (B) Scatter plots illustrating the association between biomarker levels and the MRI-assessed cortical thickness. Red lines with grey bands indicate linear regression fits with 95% confidence intervals. NPQ represents the log2-transformation of normalized target counts. N status was determined based on the surface-area weighted average cortical thickness, with < 2.7 as N+.

Additionally, 13 biomarkers—BDNF, CXCL8, CHI3L1, YWHAG, TIMP3, CCL17, IL6, VEGFD, CCL13, IL7, CD40LG, BASP1, and MSLN—showed marginally significant correlations with cortical thickness (Figure 3B). All were negatively associated with cortical thickness, except for BASP1, which showed a positive correlation.

### 3.5 Stratified biomarker association with A and N statuses

To investigate whether the association between NULISAseq biomarkers and A/N status varies across different pathological states, we evaluated the association after stratifying participants based on their A and N status. We first assessed the association of NULISAseq biomarkers with Aβ PET positivity after stratifying by N status (Figure 4A). In both stratified comparisons (A-N- vs. A+N-, A-N+ vs. A+N+), p- tau217 was the only biomarker showing a significant association, with *p*-values < 0.001 in both cases. However, several biomarkers showed marginal significance in the stratified comparisons. In the N- subgroup, ten biomarkers—CCL2, CALB2, NGF, GFAP, MAPT, p-tau231, TAFA5, CXCL10, ACHE, and NPY—showed marginally significant associations with A status. In the N+ subgroup, six biomarkers— GFAP, CRP, p-tau231, MME, NRGN, and S100A12—showed marginally significant associations with A status. A number of these biomarkers showed differential associations with A status in the N- vs. N+ subgroups. Among these biomarkers, CCL2, CALB2, NGF, MAPT, TAFA5, CXCL10, ACHE, and NPY showed stronger significance in the N- subgroup, while GFAP, p-tau231, MME, CRP, and S100A12 showed stronger associations with A status in the N+ subgroup (Supplementary Table S2).

**Figure 4.**
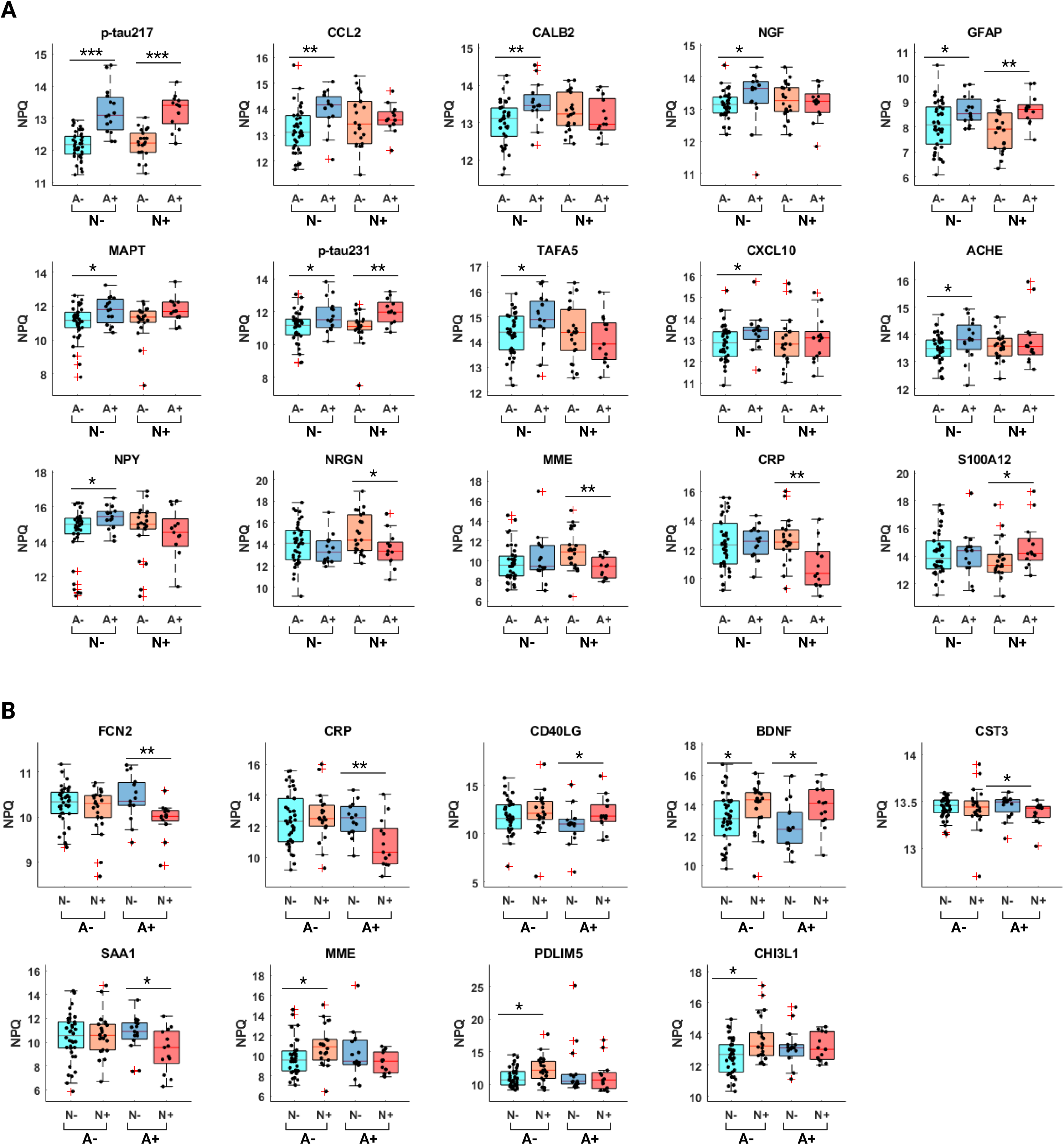
Stratified analysis for the association of NULISAseq biomarkers with A and N status. (A) Association of NULISAseq biomarkers with Aβ PET positivity, stratified by N status. (B) Association of NULISAseq biomarkers with N status, stratified by A status. Statistical significance was determined using Wilcoxon’s rank-sum test and is denoted as follows: *p < 0.05, **p < 0.01, ***p < 0.001. NPQ represents the log2-transformation of normalized target counts.

We next evaluated the association of NULISAseq biomarkers with N status, stratifying participants by their A status (Figure 4B). BDNF showed similar marginal significance in comparisons within both subgroups. PDLIM5, MME, and CHI3L1 showed a stronger association with N status within the A- subgroup, while FCN2, CRP, CD40LG, CST3, and SAA1 showed a stronger association within the A+ subgroup (Supplementary Table S3).

### 3.6 Biomarker variations across age, sex, *APOE* genotype and self-identified racial identity groups

Three biomarkers—GFAP, NEFL, and SFTPD—showed significant Spearman correlations with age. The levels of all three biomarkers increased with age, with *rho* values of 0.558 (*p* < 0.0001), 0.386 (*p* = 0.0002), and 0.372 (*p* = 0.0004), respectively (Figure 5A). Additionally, SOD1, NEFH, MDH1, FOLR1, p-tau217, PGK1, MME, IL33, VSNL1, PTN, and UCHL1 showed marginally significant correlations with age. SOD1, MDH1, PGK1, MME, and VSNL1 levels decreased with age, with the rest increasing with age.

**Figure 5.**
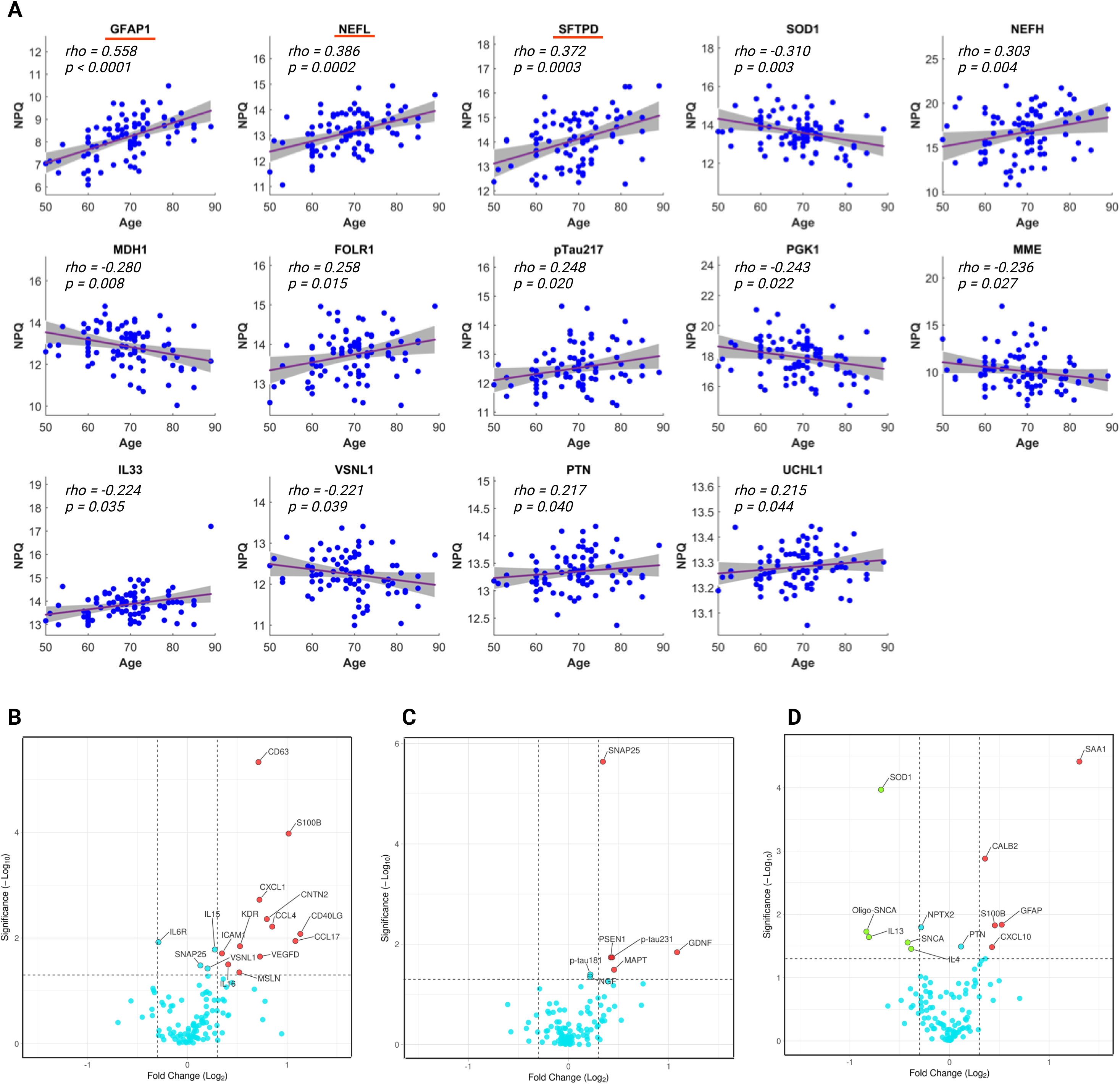
Association of NULISAseq biomarkers with age, race, *APOE* genotype, and sex. (A) Scatter plots illustrating the correlation between NULISAseq biomarkers and age. Purple lines with grey bands indicate linear regression fits with 95% confidence intervals. Rho and p values were determined using Spearman rank-based correlation. Biomarkers with red underlines indicate having significant association according to a 5% FDR cutoff. (B-D) Volcano plots of -log10(p-value) versus log2(fold change) illustrating the association between NULISAseq biomarkers with race (B), *APOE* ε4 carrier status (C), and sex (D). Up-regulated biomarkers are indicated in red, while down-regulated biomarkers are indicated in green, using a cutoff of | Log2(fold change) | > 0.3. The fold changes are based on comparing Black/African American over non-Hispanic Whites, *APOE* ε4 carrier over non-carrier, and male over female.

Self-identification as a Black/African American was associated with significantly higher levels of CD63 (*p* < 0.0001) and S100B (*p* = 0.0001) compared with non-Hispanic Whites. CD63 showed an average increase of 63%, and S100B 102% in African Americans compared with non-Hispanic Whites (Figure 5B; Supplementary Figure S1). Additionally, 14 biomarkers—CXCL1, CNTN2, CCL4, CD40LG, CCL17, IL6R, KDR, IL15, ICAM1, VEGFD, IL16, SNAP25, VSNL1, and MSLN—exhibited marginally significant differential abundance between the two groups. All except IL6R showed higher levels in African Americans.

SNAP25 showed significantly higher levels in *APOE* ε4 carriers, with a *p*-value < 0.0001 and an average increase of 27% (Figure 5C; Supplementary Figure S2). GDNF, PSEN1, p-tau231, MAPT, p- tau181, and NGF showed marginally significant differential abundance between *APOE* ε4 carriers and non-carriers. All of these proteins exhibited higher levels in *APOE* ε4 carriers.

SAA1 and SOD1 exhibited significant sex differential abundance (Figure 5D; Supplementary Figure S3). The average abundance of SAA1 was 148% higher in males, with a rank-sum *p*-value < 0.0001. In contrast, SOD1 was, on average, 38% lower in males, with a *p*-value of 0.0001. Additionally, ten biomarkers—CALB1, GFAP, S100B, NPTX2, oligomeric SNCA (oligo-SNCA), IL13, SNCA, PTN, CXCL10, and IL4—showed marginally significant sex differential abundance (Figure 5D; Supplementary Figure S3). CALB1, GFAP, S100B, PTN, and CXCL10 had higher levels in males, while the rest were higher in females.

## 4. DISCUSSIONS

Precision medicine approaches, personalizing AD interventions, will require a comprehensive panel of biomarkers that can serve as surrogate metrics for both core A, T and N pathological processes as well as other emerging pathological processes, such as cerebrovascular dysregulation, neuroinflammation, synucleinopathy, and synaptic dysfunction. A multiplex assay that can incorporate the measurement of all these biomarkers in a single assay can greatly increase assay turn-around time, reduce costs and sample volume, and minimize preanalytical errors and variation. However, multiplex proteomic assays in human plasma samples face significant challenges, including the vast dynamic range of protein concentrations, interference, cross-reactivity with other proteins and the low abundance of clinically relevant target analytes. Although several emerging high-multiplex proteomic technologies— such as antibody arrays, PLA, proximity extension assay (PEA), microsphere bead capture technology by Luminex, slow off-rate modified aptamer assay (SOMAscan), and mass spectrometry assays—can simultaneously measure hundreds to thousands of plasma proteins,^31^ it is challenging to provide comprehensive coverage of low-abundant AD blood biomarkers in a single assay.

Therefore, the availability of the NULISAseq CNS disease panel has been of great interest to the AD biomarker field. This panel utilizes a small volume of plasma samples (∼10 μl) to provide a comprehensive profile of ∼120 key proteins associated with neurodegenerative diseases. Since its advent, several studies, including one of our own, have evaluated its performance in neurodegenerative diseases, including three in AD and one in amyotrophic lateral sclerosis (ALS). ^22,23,32–34^ The findings support its alignment with established platforms for core AD biomarkers and its potential in predicting AD pathologies and monitoring ALS treatment response.^22,23,32–34^ However, further evaluation in more diverse cohorts would be helpful to validate the robustness and generalizability of the NULISAseq CNS disease panel.

Unlike our previous study in the Monongahela Youghiogheny Healthy Aging Team Neuroimaging (MYHAT-NI) cohort which consists mostly of non-Hispanic White participants, the HCP cohort evaluated herein was designed to have equal representation of Black/African Americans and non-Hispanic whites.^24^ In the present study cohort, 32 out of 88 participants self-reported as Black/African American. MYHAT-NI participants were recruited from the Rust Belt area of southwestern Pennsylvania, while HCP participants were mostly from the Pittsburgh metropolitan area. Compared with MYHAT-NI, the HCP cohort tends to be younger (median age of 70 compared with 76 for MYHAT-NI at baseline visit), with higher education levels (mean 15.7 years of education compared to 13.7), and a higher prevalence of *APOE* ε4 carriers (35.5% compared to 19.4%).

Similar to our previous findings,^22^ GFAP and NEFL exhibited strong concordance with measurements from Simoa assays using Quanterix commercial kits. This strong concordance could be attributed to the fact that GFAP and NEFL are present in higher abundance in plasma compared to other biomarkers included in the inter-platform comparison. In general, phosphorylated tau, including p-tau217, p-tau181, and p-tau231, exhibited higher concordance between NULISA assays and Simoa assays than the Aβ peptides. It is unclear whether this is due to the better stability of phosphorylated tau in plasma than Aβ peptides and, thus less prone to pre-analytical variation. Notably, while the NULISA measurements of IL6 and TNF aligned well with results from Quest immunoassays, the findings obtained from Simoa did not correlate as well with those from either platform. It is also worth noting that while several IL6 measurements from Quest assays fell around the detection limit (1.4 pg/ml), NULISA results provided better measurement resolution of these low-abundant samples, suggesting that the NULISA assay may have superior sensitivity than the Quest assay.

A total of 14 NULISAseq biomarkers showed significant associations with Aβ pathology, as assessed using dichotomized Aβ PET status or continuous Aβ accumulation according to global PiB SUVR. Consistent with their demonstrated association with brain amyloidosis, phosphorylated tau (p- tau217, p-tau231, p-tau181), GFAP, and NEFL, were on the list. Three chemokines, CCL2, CXCL10, and CXCL8, also exhibited Aβ status-dependent differential abundance, with all three being elevated in Aβ+ participants. This trend differed from what we observed previously in the MYHAT-NI cohort, where several chemokines were less abundant in Aβ+ participants or correlated with a lower accumulation rate change. This discrepancy may reflect different stages of Aβ+ participants in the HCP versus MYHAT-NI cohorts and the dynamic involvement of these chemokines in brain amyloidosis.

Chitotriosidase (CHIT1, chitinase 1), a potential biomarker for microglial activation with a role in regulating microglial polarization and Aβ oligomer proteostasis,^35^ was also elevated in A+ participants. Neurogranin (NRGN), a post-synaptic protein involved in memory formation, was decreased in A+ participants. Several publications have documented elevated CSF NRGN levels in AD patients ^36–40^. However, NRGN was found in lower abundance in plasma exosomes derived from AD and MCI patients than healthy controls.^40–42^ These findings suggest that NRGN may have different dynamics in CSF and plasma, underscoring the importance of considering both CSF and plasma measurements to comprehensively understand its role in AD pathology.

The NULISAseq CNS Panel 120 showed a weaker association with N status than Aβ status, with only marginal significance observed for 14 biomarkers. Among the most significant were the neurotrophic factor BDNF, tissue inhibitor of metalloproteinase-3 (TIMP3), and ficolin-2 (FCN2). BDNF and TIMP3 were elevated in N+ participants, while FCN2 was decreased. Intriguingly, in our previous work with the MYHAT-NI cohort, BDNF and TIMP3 were decreased in A+ participants, but no N-status-dependent changes were observed.^22^ These findings underscore the complexity of the dynamic interplay between these biomarkers and different AD pathophysiological processes, necessitating comprehensive longitudinal studies to understand the temporal dynamics and causal relationships of these biomarkers. Additional biomarkers with marginally significant associations with N status included six cytokines: CCL13, CCL17, CXCL8, CD40LG, IL6, and IL7, all negatively associated with MRI-derived cortical thickness.

Findings from stratified analysis of the association of NULISAseq biomarkers with amyloidosis and neurodegeneration highlight the complex interplay between these two aspects of Alzheimer’s disease. Tau (MAPT) and phosphorylated tau generally showed similar association with Aβ PET status, independently of the neurodegeneration status. Interestingly both CRP and FCN2 showed much lower abundance in the A+N+ participants, but similar levels in A-N-, A-N+, and A-N+, suggesting that the presence of both Aβ pathology and neurodegeneration may be necessary to observe significant changes in these biomarkers, indicating a potential synergistic effect.

Several NULISAseq biomarkers demonstrated significant associations with common AD risk factors. For instance, GFAP, NEFL, and SFTPD significantly correlated with age; these findings for GFAP and NEFL are in agreement with documented literature evidence.^43–45^ CD63 and S100B exhibited significant differential abundance between Black/African Americans and non-Hispanic Whites. SNAP25, a biomarker for synaptic degeneration, showed a significant *APOE* ε4-dependent abundance change. SAA1 is an acute-phase protein and has been suggested to have a potential role in exacerbating neuronal inflammation.^46^ It was found to be significantly higher in males in our study. SOD1, an essential antioxidant enzyme with brain-protective functions,^47–49^ was, on average, lower in males. Additionally, several biomarkers showed marginal significance with these risk factors. These findings underscore the importance of considering potential confounding factors and other coexisting morbidities when interpreting the significance of these biomarkers with AD pathologies.

Our study’s strengths include the application of a novel multiplex and neurodegenerative disease-targeting immunoassay to a community-based, racially diverse, preclinical Alzheimer’s disease cohort. This allowed us to evaluate the potential of the novel technology in providing comprehensive proteomic profiling of pathophysiological status. The small volume requirement of this technology could help streamline the implementation of blood-based biomarker tests in resource-limited settings. However, our study’s limitations include the relatively small sample size and the lack of long-term follow-up, which limits the ability to assess the efficacy of the NULISAseq CNS Disease Panel 120 in personalized AD management.

Importantly, we have demonstrated that several novel plasma biomarkers – beyond Aβ, tau, p-tau, GFAP and NEFL – reveal biological changes in processes such as synaptic function, microglial activation, peripheral inflammation, and oxidative stress that define the pathophysiological state of individuals at different stages of Aβ pathology and neurodegeneration that characterize Alzheimer’s disease.

## Data Availability

All data produced in the present study are available upon reasonable request to the authors, provided requests are in accordance with the terms of the IRB approval and the laws of the Commonwealth of Pennsylvania and the United Sttes.

## ACKNOWLEDGEMENTS/CONFLICTS/FUNDING SOURCES/CONSENT STATEMENT

## Acknowledgements

We thank the HCP study participants and their families and caregivers.

## Consent statement

All participants provided written consent, and the University of Pittsburgh Institutional Review Board approved the study.

## HIGHLIGHTS

- Classical AD biomarkers measured by NULISAseq CNS panels showed strong concordance with those measured using established immunoassay methods from Quanterix and Quest, with GFAP and NfL exhibiting the strongest correlation.
- NULISAseq proteomic analysis identified several plasma biomarkers strongly associated with AD pathology in a biracial community cohort of older adults. Notably, p-tau217, GFAP, and p-tau231 displayed the strongest association with Aβ pathology, while BDNF was strongly associated with neurodegeneration.
- We demonstrate that plasma biomarker levels could be influenced by age, sex, *APOE* genotype, and self-identified race. Specifically, GFAP, NfL, and SFTPD showed a strong association with age; CD63 and S100B with self-identified race; SNAP25 with APOE genotype; and SAA1 and SOD1 with significant sex differences.

## RESEARCH IN CONTEXT

**1. Systematic review:** The authors reviewed the current state of blood-based biomarker assays for Alzheimer’s disease (AD) detection using PubMed. While many promising blood biomarkers have been linked to classical AD pathologies, there is a lack of biomarkers for co-pathologies such as inflammation, cerebrovascular lesions, and synaptic dysfunction. A multiplex assay capable of providing biological evidence of various AD pathologies is urgently needed to improve the clinical management of AD.
**2. Interpretation:** Using the HCP community-based biracial preclinical cohort, this study evaluated the utility of the NULISAseq CNS panel, an innovative multiplex immunoassay targeting approximately 120 key proteins associated with neurodegenerative diseases. Our findings demonstrate robust concordance between NULISAseq measurements and those obtained with established technologies for classical AD biomarkers. Additionally, we uncovered new candidate biomarkers for amyloid pathology and neurodegeneration.
**3. Future directions:** Further work involving large cohorts with longitudinal follow-up is needed to confirm the association of novel biomarkers with AD pathologies and to validate the utility of the NULISAseq CNS panel in personalized AD management.

