## Supplementary Figure S1 for "Novel plasma biomarkers of amyloid plaque pathology and cortical thickness: evaluation of the NULISA targeted proteomic platform in an ethnically diverse cohort"

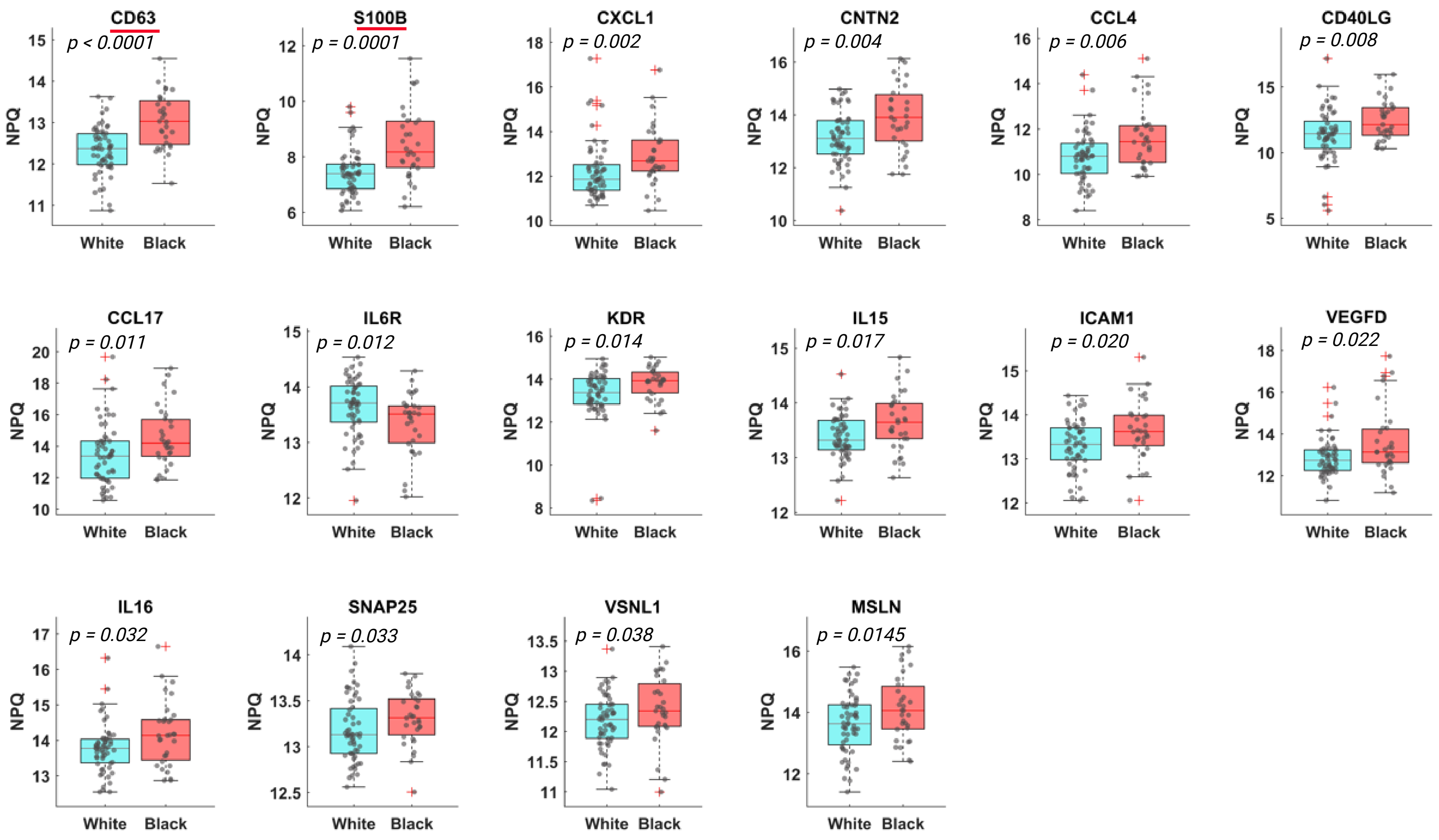


**Figure S1.** Box plot distributions of NULISAseq biomarkers with significant or marginal significant racial differences. P values were determined using Wilcoxon's rank-sum test.
