## Supplementary Figure S2 for "Novel plasma biomarkers of amyloid plaque pathology and cortical thickness: evaluation of the NULISA targeted proteomic platform in an ethnically diverse cohort"

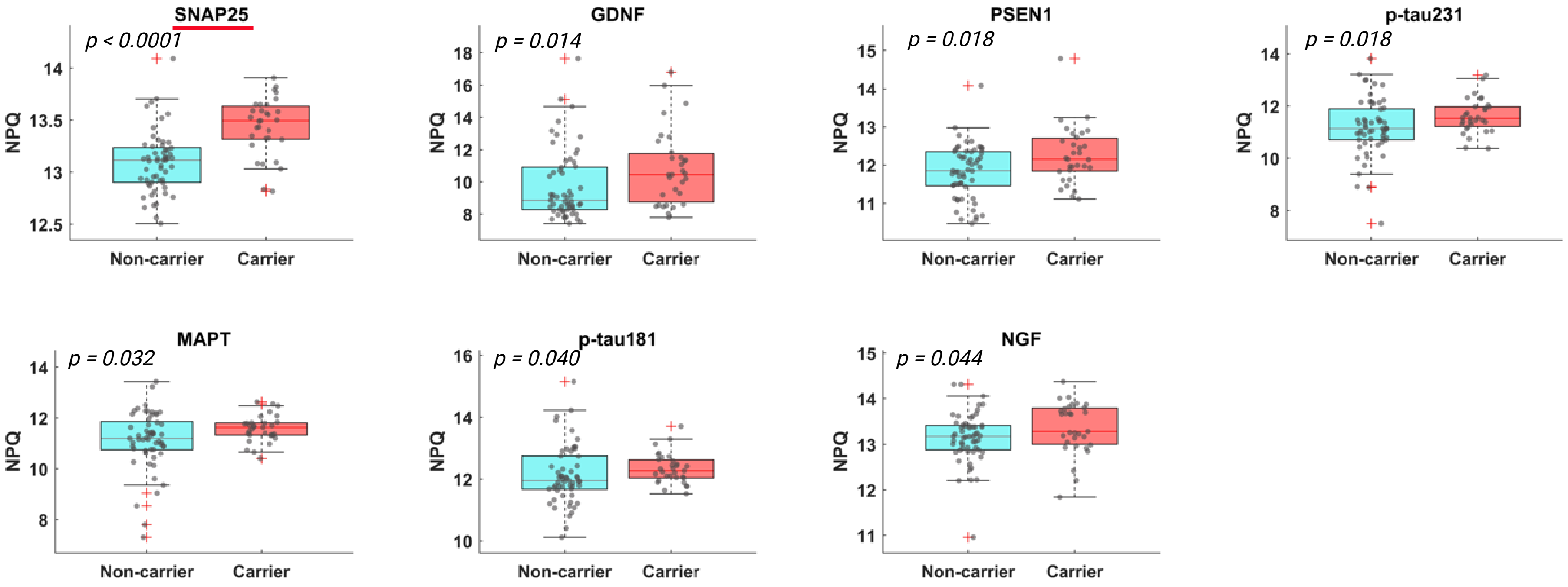


**Figure S2.** Box plot distributions for NULISAseq biomarkers with significant or marginal significant differential abundance in APOE ε4 carriers vs. non-carriers. P values were determined using Wilcoxon's rank-sum test.
