## Supplementary Figure S3 for "Novel plasma biomarkers of amyloid plaque pathology and cortical thickness: evaluation of the NULISA targeted proteomic platform in an ethnically diverse cohort"

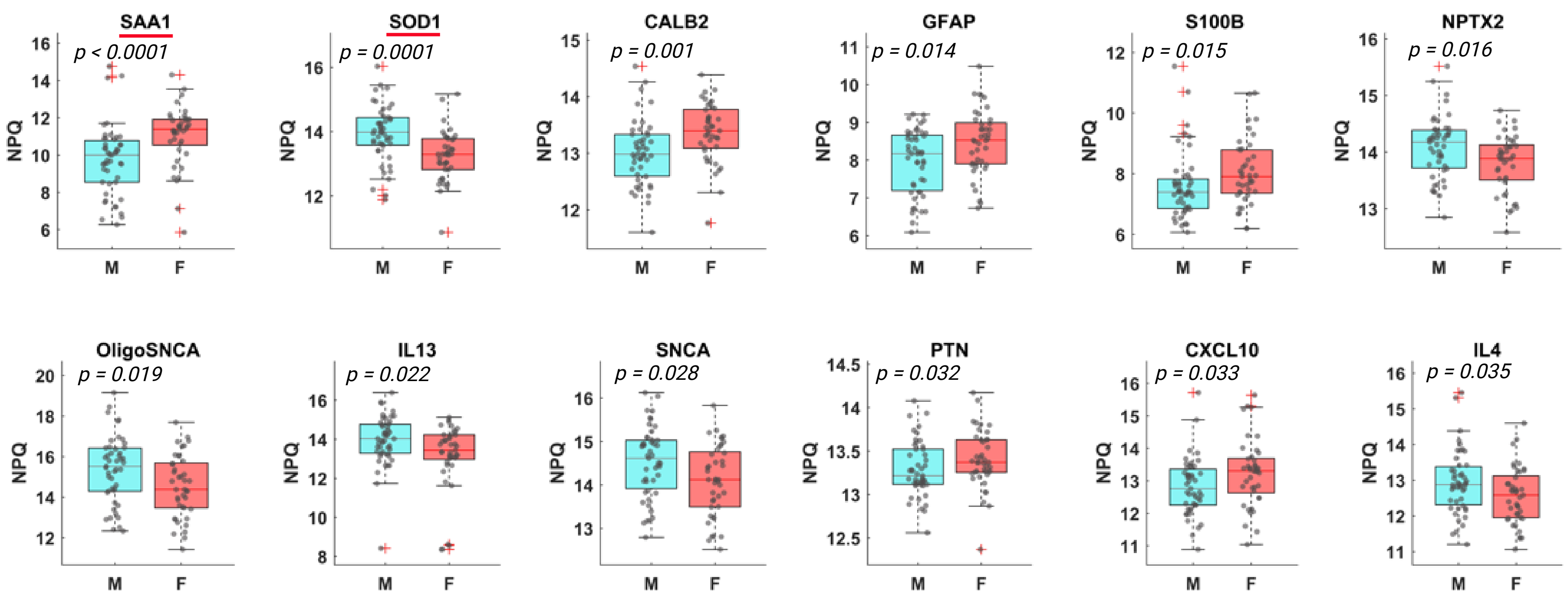


**Figure S3.** Box plot distributions for NULISAseq biomarkers with significant or marginal significant sex differences. P values were determined using Wilcoxon's rank-sum test.
