## Supplementary Table S1 for "Novel plasma biomarkers of amyloid plaque pathology and cortical thickness: evaluation of the NULISA targeted proteomic platform in an ethnically diverse cohort"

**Supplementary Table S1:** Impact of common risk factors on the significance of the association between NULISAseq biomarkers and amyloid pathology.

|  |  |  | **Logistic Regression Model** | | | | |  |
| --- | --- | --- | --- | --- | --- | --- | --- | --- |
| **Biomarker** | **Rank-sum** | **No covariate** | **+ Age** | **+ Race** | **+ APOE** | **+ Sex** | **+ Education** | **+ Sex + Age + APOE** |
| **p-tau217** | <0.0001* | <0.0001* | <0.0001* | <0.0001* | <0.0001* | <0.0001* | <0.0001* | <0.0001* |
| **GFAP** | 0.0004* | 0.0013* | 0.0270 | 0.0014 | 0.0010* | 0.0003* | 0.0030 | 0.0068 |
| **p-tau231** | 0.0005* | 0.0007* | 0.0009 | 0.0003* | 0.0008* | 0.0010* | 0.0010 | 0.0018 |
| **MAPT** | 0.0039 | 0.0046 | 0.0033 | 0.0025 | 0.0048 | 0.0049 | 0.0045 | 0.0040 |
| **CCL2** | 0.0064 | 0.0194 | 0.0210 | 0.0106 | 0.0183 | 0.0169 | 0.0192 | 0.0227 |
| **p-tau181** | 0.0187 | 0.0159 | 0.0172 | 0.0060 | 0.0171 | 0.0207 | 0.0183 | 0.0298 |
| **CHIT1** | 0.0210 | 0.0644 | 0.1435 | 0.0559 | 0.0603 | 0.0571 | 0.0563 | 0.1457 |
| **CXCL8** | 0.0305 | 0.1058 | 0.2062 | 0.0765 | 0.0957 | 0.1041 | 0.0737 | 0.2248 |
| **NRGN** | 0.0326 | 0.0466 | 0.0720 | 0.0652 | 0.0376 | 0.0483 | 0.1382 | 0.0796 |
| **NEFL** | 0.0407 | 0.0394 | 0.3930 | 0.0249 | 0.0277 | 0.0303 | 0.1451 | 0.3316 |

Note: * indicates false discovery rates < 0.05.
