## Supplementary Table S2 for "Novel plasma biomarkers of amyloid plaque pathology and cortical thickness: evaluation of the NULISA targeted proteomic platform in an ethnically diverse cohort"

**Supplementary Table S2:** Association of NULISAseq biomarkers with A status stratified by N status.

| **Biomarker** | **N-** | | **N+** | |
| --- | --- | --- | --- | --- |
|  | **p-value** | **Fold change** | **p-value** | **Fold change** |
| p-tau217 | <0.0001 | 2.11 | <0.0001 | 2.02 |
| GFAP | 0.0322 | 1.49 | 0.0018 | 1.89 |
| CRP | 0.7297 | 1.03 | 0.0060 | 0.30 |
| p-tau231 | 0.0355 | 1.64 | 0.0066 | 1.93 |
| MME | 0.5083 | 1.44 | 0.0066 | 0.32 |
| NRGN | 0.2401 | 0.69 | 0.0458 | 0.38 |
| S100A12 | 0.7746 | 0.99 | 0.0458 | 1.95 |
| MAPT | 0.0338 | 1.63 | 0.0787 | 1.53 |
| NPY | 0.0472 | 1.77 | 0.3478 | 0.87 |
| CALB2 | 0.0059 | 1.41 | 0.4424 | 0.92 |
| TAFA5 | 0.0391 | 1.47 | 0.6946 | 0.84 |
| NGF | 0.0214 | 1.18 | 0.7457 | 0.93 |
| CCL2 | 0.0018 | 1.74 | 0.8244 | 1.03 |
| CXCL10 | 0.0430 | 1.39 | 0.9049 | 1.07 |
| ACHE | 0.0451 | 1.26 | 0.9049 | 1.19 |
