## Supplementary Table S3 for "Novel plasma biomarkers of amyloid plaque pathology and cortical thickness: evaluation of the NULISA targeted proteomic platform in an ethnically diverse cohort"

**Supplementary Table S3:** Association of NULISAseq biomarkers with N status stratified by A status.

| **Biomarker** | **A-** | | **A+** | |
| --- | --- | --- | --- | --- |
|  | **p-value** | **Fold change** | **p-value** | **Fold change** |
| MME | 0.0102 | 2.24 | 0.3334 | 0.50 |
| PDLIM5 | 0.0132 | 2.41 | 0.7822 | 0.56 |
| CHI3L1 | 0.0171 | 2.07 | 1.0000 | 1.03 |
| BDNF | 0.0470 | 1.75 | 0.0304 | 2.39 |
| CD40LG | 0.1746 | 1.39 | 0.0270 | 2.23 |
| CRP | 0.4569 | 1.16 | 0.0099 | 0.34 |
| FCN2 | 0.5144 | 0.92 | 0.0066 | 0.72 |
| CST3 | 0.8240 | 1.00 | 0.0304 | 0.95 |
| SAA1 | 0.8841 | 1.19 | 0.0382 | 0.38 |
